# Moonlit Misconceptions: A South Florida Veterans’ Hospital Perspective on Full Moon Emergency Room Visits

**DOI:** 10.1101/2024.10.22.24315836

**Authors:** David J. VanMeter, Timothy S. Strebel, Stephen M. Mastorides, Andrew A. Borkowski

## Abstract

The belief that the full moon influences human behavior and health outcomes has persisted for centuries despite limited scientific evidence. This study examined whether there is a correlation between the lunar cycle and emergency department (ED) visits and hospital admissions among veterans in South Florida from 2013-2023. Data on 465,838 ED visits resulting in 110,640 admissions and moon illumination fraction data were analyzed. The results showed no significant difference in the mean number of ED visits, admissions, or percentage of visits resulting in admission across different moon illumination fractions, including full moons. While some past studies have suggested links between the lunar cycle and health conditions like seizures and psychiatric issues, this study found no association between the full moon and ED utilization patterns in the South Florida veteran population over an 11-year period. The findings contradict the long-standing belief about the lunar effect on hospital services and do not support adapting staffing or resources based on the moon’s phase.

## 1. INTRODUCTION

The enduring belief in the moon’s potential to influence human behavior and health outcomes, a belief that has spanned centuries, continues to captivate despite the dearth of scientific evidence to substantiate such assertions. One area that has piqued particular interest is the so-called ‘lunar effect’ on hospital admissions and emergency department (ED) visits.

Despite numerous studies exploring the potential link between the full moon and increased ED visits, the evidence remains inconclusive. For instance, a 1996 study concluded that a full moon did not affect ED patient volume, ambulance runs, admissions, or admissions to a monitored unit.^1^ An additional 2005 study found no significant difference in the number of overnight psychiatric ED visits on full moon nights compared to non-full moon nights.^2^

However, it is worth noting that other studies have hinted at a potential connection. A 2020 study, for example, found that admissions of schizophrenia patients increased most during the first-quarter lunar phases, with the full moon exerting some influence.^3^ In addition, a 2023 study focused specifically on the effect of ‘supermoons’ (perigee full moons) on emergency department behavioral health volume and total volume. It discovered a statistically significant higher percentage of behavioral health volume as a percent of total volume on supermoon days compared to non-supermoon days. However, the overall difference in total patient volume was not statistically significant, indicating a complex relationship between lunar phases and health outcomes.^4^

Few other studies have suggested the mild effect of the full moon on increased time in ambulance transportation for road crashes and increased gastrointestinal bleeding.^5,6^

As there is no existing research on the correlation between the full moon, emergency department visits, and hospital admissions among South Florida’s veteran population, we have decided to undertake a study to address this knowledge gap. We aimed to investigate whether there is a significant increase in the number of ED visits and hospital admissions during full moon periods compared to other times of the lunar cycle. Through this study, we hoped to gain a better understanding of the potential impact of the lunar cycle on healthcare utilization among veterans in our region.

## 2. MATERIALS AND METHODS

The data for the ED visits and admissions were extracted from the James A. Haley Veterans’ Hospital Corporate Data Warehouse for the period from January 1, 2013, to December 31, 2023. Our dataset did not contain any patient-identifiable information. During the eleven years, there were 465,838 ED visits that resulted in 110,640 admissions. The average number of ED visits was 116 (minimum 22, maximum 196, standard deviation 28.63), and the average number of ED admissions was 27.54 (minimum 4, maximum 53, standard deviation of 7.44). In addition, we obtained the Fraction of the Moon Illuminated data for each day from the Astronomical Applications Department of the U.S. Naval Observatory for each day of the same period.^7^ There were 161 occurrences of the full moons during the investigated eleven-year period.

To perform statistical analysis, we used the following Python libraries: Pandas, Numpy, SciPy, StatsModels and Altair. The primary unit of analysis was the calendar day of observed Emergency Department activity. We analyzed the fraction of the moon illuminated for each day as our independent variable and three response variables: the number of ED visits, number of ED visits resulting in admissions, and percentage of ED visits resulting in admissions. We used both continuous and categorical representations of the fraction of the moon illuminated by dividing our dataset into six parts: 0 (new moon), 0.1-0.24, 0.25-0.49, 0.50-0.74, 0.75-0.99, and 1 (full moon).

Our statistical analysis consisted of two methods. First, we conducted correlation tests to identify potential relationships between the continuous value of fraction of the moon and our three response variables. The purpose of these tests was to assess whether there was a general relationship between the moon cycle and our three response variables. Prior to conducting this test, we examined the distributional characteristics of our data using QQ-plots and statistical control charts to assess the appropriate statistical test. We determined that the Spearman Rank Correlation test was most appropriate. We reported the correlation coefficient and two-sided significance values of the tests.

Second, a cross-sectional statistical comparison of estimates of the population mean derived from our sample data from the full moon category to the remaining five of categories of fraction of the moon across two of our three response variables. The purpose of using comparison tests is to try and identify significant differences to our responses during the full moon cycle. We left out the percentage of ED visits resulting in admission due to its inappropriate use as a percentage for a comparison test. We observed the shape of our sample data using QQ-plots to ascertain the appropriate statistical comparison test. We decided to use the two-sided Student’s T-test and report the T-statistic and significance values for each comparison.

## 3. RESULTS

The mean and standard deviation numbers for ED visits, ED admissions, and percentage of ED visits resulting in admissions, were similar for all the six moon fractions, including the full moon. Table 1, Figure 1. Pearson Correlation Coefficient (r) was close to zero when comparing six fractions of the moon illumination to the number of ED visits (Pearsons R: -0.005688, P-value 0.7185), number of ED visits resulting in admissions (Pearsons R: -0.005482, P-value 0.7283), and percentage of ED visits resulting in admissions (Pearsons R: -0.007636, P-value 0.6285). Figure 2. A two-sided Student’s T-test comparing the full moon fraction to other fractions of moon illumination, in reference to the number of ED visits and later to the number of ED admissions, showed no statistically significant differences. Table 2.

**Table 1.** Relationship between number of ED visits, ED admissions, percentage of ED visits resulting in admissions, and the fractions of the moon illuminated.

| Fraction of the Moon Illuminated |  | 0 (New Moon) | 0.1-0.24 | 0.25-0.49 | 0.50-0.74 | 0.75-0.99 | 1 (Full Moon) |
| --- | --- | --- | --- | --- | --- | --- | --- |
| ED Visits | Mean | 117.286585 | 115.934952 | 116.04246 | 116.701625 | 115.389313 | 115.664596 |
|  | Standard Deviation | 29.814511 | 28.369892 | 29.071501 | 29.044185 | 28.305517 | 28.211909 |
| ED Admissions | Mean | 28.256098 | 27.696444 | 27.04978 | 27.503693 | 27.529262 | 28.074534 |
|  | Standard Deviation | 6.965843 | 7.332017 | 7.467653 | 7.77563 | 7.460692 | 6.844298 |
| Percentage of ED Visits Resulting in Admissions | Mean | 0.246133 | 0.242538 | 0.236368 | 0.238128 | 0.241488 | 0.247448 |
|  | Standard Deviation | 0.048883 | 0.049157 | 0.048113 | 0.048173 | 0.047813 | 0.046688 |

**Table 2.** A two-sided Student’s T-test comparing the full moon fraction to other fractions of moon illuminated regarding the number of ED visits and to the number of ED admissions.

| Fraction of the Moon Illuminated |  | 0 (New Moon) | 0.1-0.24 | 0.25-0.49 | 0.50-0.74 | 0.75-0.99 | 1 (Full Moon) |
| --- | --- | --- | --- | --- | --- | --- | --- |
| ED Visits | T-Statistic | 0.5036 | 0.1133 | 0.1492 | 0.4094 | -0.1158 | 0 |
|  | P-Value | 0.6149 | 0.9098 | 0.8814 | 0.6823 | 0.9078 | 1 |
| ED Admissions | T-Statistic | 0.237 | -0.6178 | -1.591 | -0.8559 | -0.8782 | 0 |
|  | P-Value | 0.8128 | 0.5368 | 0.1121 | 0.3923 | 0.38 | 1 |

**Figure 1.**
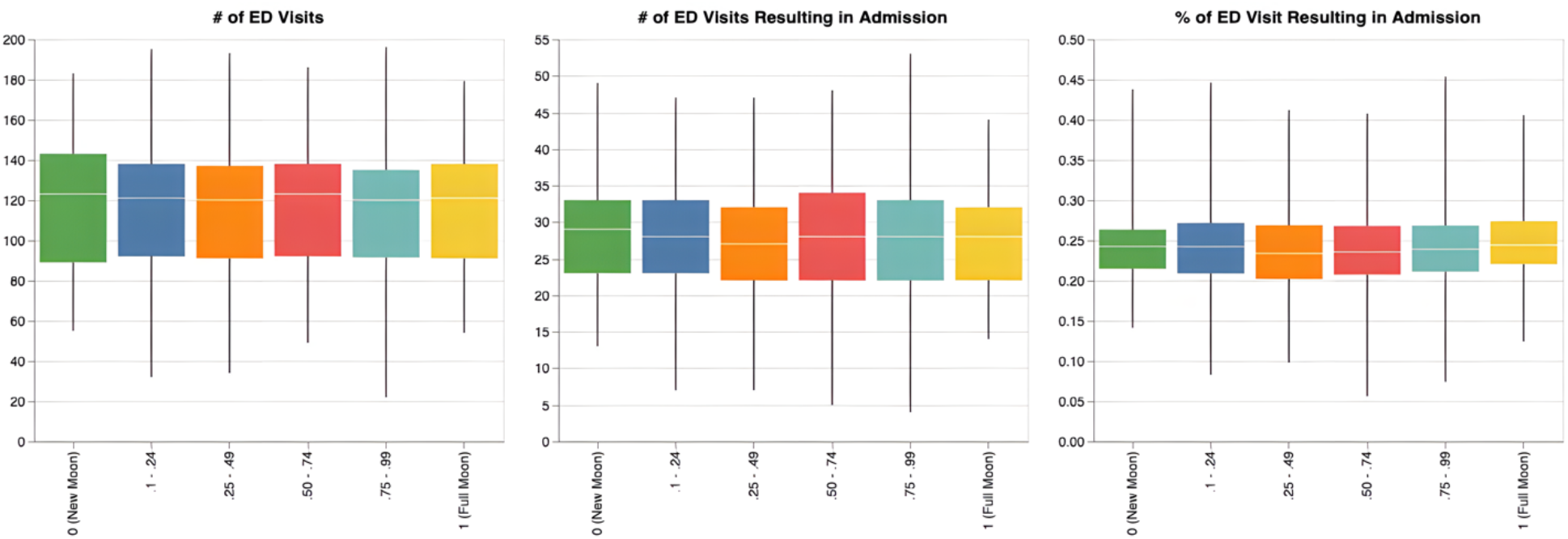
Boxplot with the relationship between the number of ED visits (left), ED admissions (middle), percentage of ED visits resulting in admissions (right), and the fractions of the moon illuminated.

**Figure 2.**
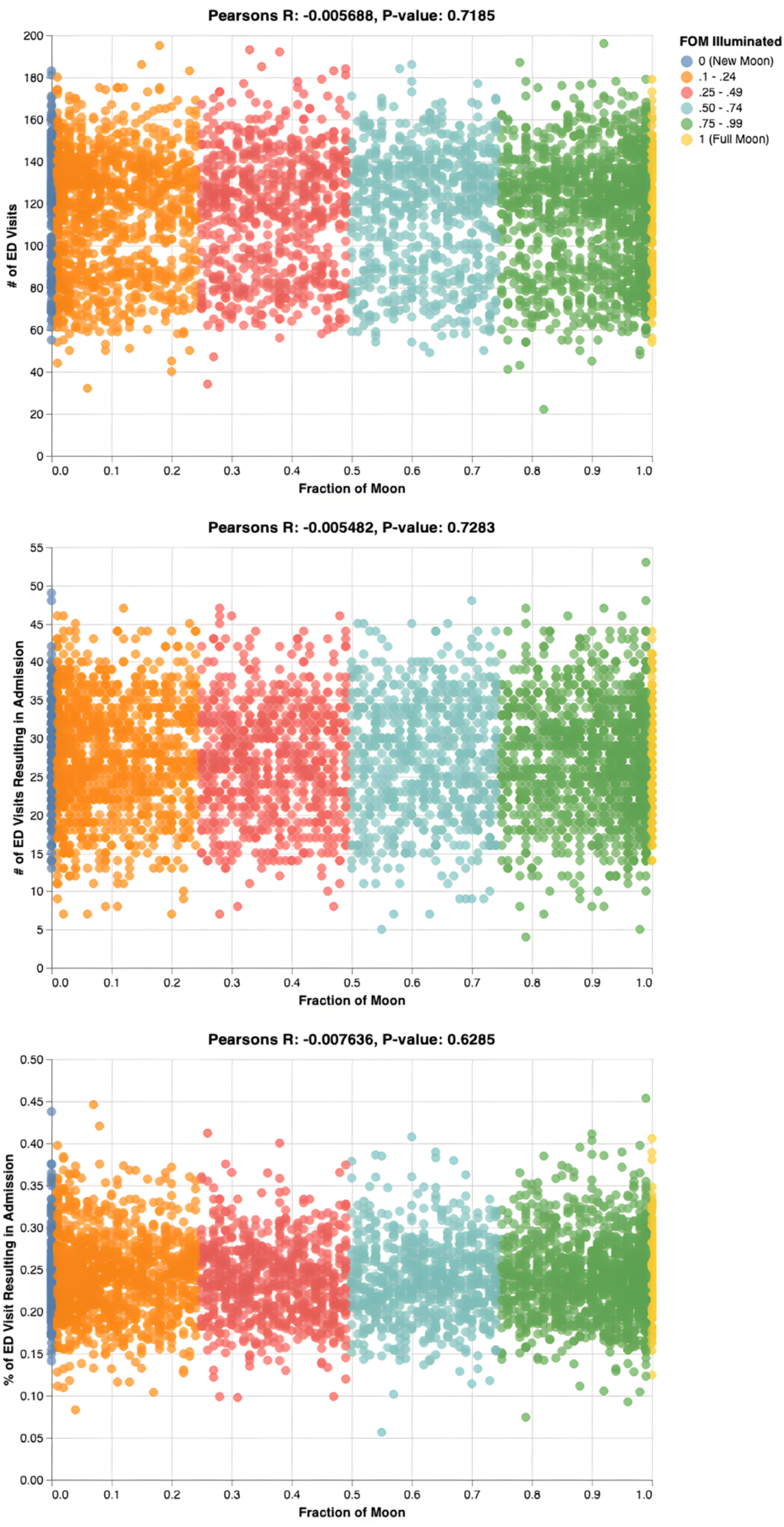
Pearson Correlation Coefficient (r) comparing six fractions of the moon illuminated to the number of ED visits (top), the number of ED visits resulting in admissions (middle), and the percentage of ED visits resulting in admissions (bottom).

## 4. DISCUSSION

We have embarked on a study investigating the correlation between the full moon, emergency department visits, and hospital admissions among South Florida’s veteran population. Since there is no existing research on this topic, we aimed to address this knowledge gap. Our goal was to determine whether there was a significant increase in the number of ED visits and hospital admissions during full moon periods compared to other times of the lunar cycle. Through this study, we hoped to understand better the potential impact of the lunar cycle on healthcare utilization among veterans in our region. The study utilized the data from January 1, 2013, to December 31, 2023, covering eleven years, during which there were 465,838 ED visits that resulted in 110,640 hospital admissions.

According to our study, there is no connection between the full moon and the number of visits or admissions to the emergency department (ED). Moreover, we have found no correlation between the full moon and the percentage of ED visits that lead to admissions, which contradicts the popular belief that patients with more severe health issues flock to the ED during full moons. We have also discovered that there is no connection between different quarters of moon illumination and ED visits, admissions, or the percentage of ED visits that result in admissions.

Over the years, various studies have investigated the full moon’s influence on various health conditions.

Several studies investigated the relationship between the full moon and ED visits. The 1996 study conducted a retrospective analysis of the hospital electronic records of all patients seen in an ED during a 4-year period in an ED of a suburban community hospital. A full moon occurred 49 times during the 4-year study period. The study found no significant differences in total patient visits, ambulance runs, admissions to the hospital, or admissions to a monitored unit on days of the full moon.^1^

Another study investigated the psychiatric ED visits on full moon nights over a five-year period. That study also showed no significant difference between the number of patients seen on full- and non-full-night nights.^2^

According to a study conducted in Henan province of China, individuals with schizophrenia were more stable during the new moon phase. However, their condition was more likely to worsen during the first quarter and the full moon phase. Patients with paranoid schizophrenia were particularly vulnerable to deterioration during the full moon phase.^3^

There is a common belief that the occurrence of seizures is linked to the full moon. While this idea has been around for centuries, recent studies have shed some light on the matter. One study looked at the neurologic records of an emergency unit between the years 1999 and 2003 to determine if there was a connection between lunar phases and seizures. The results showed that there was indeed a clustering of seizures around the full moon period, which supports the ancient belief that seizure frequency increases during full moon days.^8^ Another study, conducted over a period of three years, reviewed the occurrence of seizures in an epilepsy monitoring unit. It found no correlation between the total number of seizures, including both epileptic and non-epileptic, and the full moon. However, the study did observe an increase in non-epileptic seizure episodes during the full moon and an increase in epileptic seizures during the last quarter.^9^

A team of researchers conducted a study to investigate whether there is any correlation between lunar phases and acute myocardial infarction or sudden cardiac death. They concluded that there is no evidence to suggest that lunar phases are associated with an increased risk of these events.^10^

In yet another study, no significant differences were found between the phases of the lunar cycle and the frequency of births, delivery route, births to multigravid women, or birth complications. ^11^

It’s interesting to note that one study has shown that there’s a higher risk of fatal motorcycle crashes during a full moon period.^12^ Another study has indicated a rise in ambulance transport due to road crashes during full moon periods. This information should be considered when planning for full moon ambulance coverage.^5^

## 5. CONCLUSION

Our research has found no correlation between the full moon and the number of visits or admissions to the South Florida Veteran Emergency Department (ED) during the eleven years studied. Despite some studies that suggest otherwise, we have found no connection between different quarters of moon illumination and ED visits, admissions, or the percentage of ED visits that result in admissions.

## Data Availability

The data for the fractions of the moon illuminated is available at https://aa.usno.navy.mil/data/MoonFraction

## ACKNOWLEDGMENTS

We want to thank Scott A. Dillon for helping us acquire the data for this project. We also would like to thank Sarah Pazos of the US Naval Observatory for providing us with data on moon illumination.

This project was submitted to the James A. Haley Veterans Hospital Research and Development Committee, which includes an ethics review, and was approved as a quality assurance project.

This material results from work supported by the resources and the use of the James A. Haley Veterans’ Hospital facilities.

## REFERENCES

1. Thompson DA, Adams SL. The full moon and ED patient volumes: Unearthing a myth. Am J Emerg Med. 1996;14(2):161–164. doi:10.1016/S0735-6757(96)90124-2

2. Kung S, Mrazek DA. Psychiatric Emergency Department Visits on Full-Moon Nights. Psychiatr Serv. 2005;56(2):221–a. doi:10.1176/appi.ps.56.2.221-a

3. Wang RR, Hao Y, Guo H, et al. Lunar cycle and psychiatric hospital admissions for schizophrenia: new findings from Henan province, China. Chronobiol Int. 2020;37(3):438–449. doi:10.1080/07420528.2019.1625054

4. Drsquo A, Orazio-Bradfield Taylor J, et al. Perigee Full Moons (“Supermoons”) and its Effect on Emergency Department Behavioral Health Volume and Total Volume. Mathews J Emerg Med. 2023;8(2):1–5. doi:10.30654/MJEM.10054

5. Onozuka D, Nishimura K, Hagihara A. Full moon and traffic accident-related emergency ambulance transport: A nationwide case-crossover study. Sci Total Environ. 2018;644:801–805. doi:10.1016/j.scitotenv.2018.07.053

6. Román EM, Soriano G, Fuentes M, Gálvez ML, Fernández C. The influence of the full moon on the number of admissions related to gastrointestinal bleeding. Int J Nurs Pract. 2004;10(6):292–296. doi:10.1111/j.1440-172x.2004.00492.x

7. Fraction of the Moon Illuminated. Accessed April 25, 2024. https://aa.usno.navy.mil/data/MoonFraction

8. Polychronopoulos P, Argyriou AA, Sirrou V, et al. Lunar phases and seizure occurrence: just an ancient legend? Neurology. 2006;66(9):1442–1443. doi:10.1212/01.wnl.0000210482.75864.e8

9. Benbadis SR, Chang S, Hunter J, Wang W. The influence of the full moon on seizure frequency: myth or reality? Epilepsy Behav EB. 2004;5(4):596–597. doi:10.1016/j.yebeh.2004.04.001

10. Eisenburger P, Schreiber W, Vergeiner G, et al. Lunar phases are not related to the occurrence of acute myocardial infarction and sudden cardiac death. Resuscitation. 2003;56(2):187–189. doi:10.1016/s0300-9572(02)00298-8

11. Arliss JM, Kaplan EN, Galvin SL. The effect of the lunar cycle on frequency of births and birth complications. Am J Obstet Gynecol. 2005;192(5):1462–1464. doi:10.1016/j.ajog.2004.12.034

12. Redelmeier DA, Shafir E. The full moon and motorcycle related mortality: population based double control study. BMJ. 2017;359:j5367. doi:10.1136/bmj.j5367

